# Rapid, label-free surface plasmon resonance discrimination between Bothrops and Crotalus venoms using clinical antivenom as the capture reagent

**DOI:** 10.1101/2025.08.26.25334415

**Authors:** Ely S. F. Borges, Jomar S. Vasconcelos, Arthur A. Melo, Fernanda C. C. L. Loureiro, Karla P. O. Luna, Antonio M. N. Lima

## Abstract

Snake envenoming is recognized as a global health problem, affecting thousands of people every year. One of the main challenges in addressing this issue is the correct identification and treatment of these envenomations, mainly in locations where people don’t have easy access to hospitals. In Brazil, the genus Bothrops is responsible for the majority of envenomations, followed by Crotalus. This study reports a simple methodology for detecting raw venom from snakes of the genus Bothrops, through interaction with their corresponding antibodies, using a high-sensitivity optical biosensor. The protocol consists of adding antibodies (present in the commercial antivenom) to the sensor surface, followed by the addition of Crotalus venom (nonspecific), and then Bothrops venom (specific), resulting in changes in the refractive index, to evaluate cross reactions between them. Different concentrations of raw venom from snakes of the genus Bothrops and Crotalus were tested, starting with a concentration of 6.784μgmL^−1^ and progressing until reaching the minimum detectable concentration. The binding capacity of venom to antivenom was investigated at two concentrations of antivenom: 5 μgm^−1^ and 50 μgmL^−1^. Both antivenom and snake venoms were solubilized in phosphate buffered saline (PBS). The results show an accurate detection of the antigen of interest (Bothrops venom), tested at different concentrations. The biosensor was able to detect venom up to a concentration of 0.848 μgmL^−1^. In addition, no interference from nonspecific binding between the Bothrops antivenom and Crotalus venom was detected. The detection of specific venom (Bothrops) occurred in a satisfactory time (up to 14 minutes). The results provide evidence that the biosensor and the methodology employed can be considered a diagnostic model under development, which can help health workers to better identify and treat envenomated people.

## 1 INTRODUCTION

Snake envenoming is a serious health problem that can lead to death [1], since the toxins present in the venom of these animals can cause damage of varying magnitudes to physiological systems [2]. According to the World Health Organization (WHO) [1], 4.5 to 5.4 million people are bitten by snakes annually. Among these, 1.8 to 2.7 million develop clinical diseases and 81,000 to 138,000 die from complications. However, due to underreporting, the true extent and impacts of snakebites are still unknown [3]. In Brazil, the families Viperidae (genera *Bothrops* and *Bothrocophias, Crotalus* and *Lachesis*) and Elapidae (genera *Micrurus* and *Leptomicrurus*) comprise the venomous snakes of relevance to public health, and snakes of the genus Bothrops are responsible for the largest number of accidents [4].

Snake venoms are composed of up to 90% proteins; some families of these proteins have hundreds of isoforms [5]. Snake envenomation has the potential to trigger severe paralysis (compromising the ability to breathe), bleeding disorders, permanent kidney damage and severely damage local tissues, resulting in irreversible disabilities and limb amputations [1]. The clinical manifestations of snake envenomation vary according to the chemical composition of each venom: Envenomations by snakes of the Elapidae family usually induce neurotoxic, cytotoxic, and cardiotoxic manifestations, while envenomations by snakes of the Viperidae family usually cause myotoxicity and hemotoxicity [2].

High-quality antivenoms (which follow appropriate production standards and undergo rigorous evaluation before being tested in patients) represent the most efficient therapeutic approach to prevent most of the impacts resulting from snake envenomation and are officially recognized as essential medicines by the WHO [1] in combination with support treatment [6]. The success of antivenom therapy depends on the correct and early diagnosis of the snake involved in the envenomation, therefore, ensuring access to rapid diagnostics can promote better results [7]. The decision on the antivenom to be applied is based on the clinical characteristics and/or investigations available at the point of care, such as ancillary laboratory tests [6]. In view of this, it is crucial to develop rapid, highly reliable and specific tests to detect the type of envenomation to provide better treatment effectiveness [7, 8, 9].

The usage of biosensors has contributed to great advances in the health area, thus, there is a growing need to develop simple, sensitive and economical diagnostic tools, such as biosensors, to effectively detect diseases [10]. A progressive interest in the application of sensor technologies for snake detection has been reported [11]. Researchers have been working on developing biosensors capable of detecting snake venom. However, the application of these tests in the field remains uncertain due to obstacles such as long analysis time, non-reuse of chips, high-cost equipment and reagents, meticulous preparation of the test, and the need for improvements in detection [12, 13].

Biosensors are analytical devices that make biological signals measurable by changes generated by interactions between biological components, converting them into an electrical signal [10]. A conventional biosensor consists of the following elements: analyte, bioreceptor, transducer, electronic components, and a system for interpreting the results [14, 15].

Given the urgent need to develop highly reliable and sensitive tests capable of accurately and quickly determining the occurrence of snakebite envenomation and the type of snake that caused the accident, this research proposed to develop a simple and rapid methodology using a highly sensitive optical SPR biosensor for detecting *Bothrops erythromelas* venom (venom related to snakes of the *Bothrops* genus).

## 2 MATERIAL AND METHODS

Samples of crude venom from snakes of the species *Bothrops erythromelas* were obtained from adult specimens from the Museu Vivo Répteis da Caatinga, located in the state of Paraíba, Brazil. The venom was kept at a temperature of −20°C until use. Venom samples from snakes of the species *Crotalus durissus terrificus* were provided by the Instituto Vital Brazil, located in the state of Rio de Janeiro, Brazil. The antibothrops serum (SAB) was provided by the Health Department of the State of Pernambuco and kept refrigerated at 4°C. The antibothrops serum used is produced from a combination of venoms from five species of snakes of the *Bothrops* genus (*B. jararaca, B. alternatus, B. jararacussu, B. moojeni and B. neuwiedi*) and its composition consists of F(ab`)2 fractions of heterologous, specific and purified immunoglobulins [16]. Both the antivenom serum and the snake venoms were solubilized in phosphate buffer saline (PBS), pH 7.2, immediately before the experiments.

As for the study concentrations of Bothrops venom (venom from snakes of the Bothrops genus) and Crotalus venom (venom from snakes of the *Crotalus* genus), two concentrations were tested, considered minimum and maximum. The maximum venom concentration used in the tests described here was 6.784 μgmL^−1^. The start concentration was based on previous analyses made by the group, in which 6.784 μgmL^−1^ was the lowest venom concentration tested. Therefore, in order to achieve snake venom detections at even lower concentrations, considering that venom is found in low concentrations in body fluids [17], serial dilution were conducted in PBS (1:15,000, 1:30,000, and 1:60,000) until reaching the minimum concentration at which the biosensor detected interaction, which was 0.848 μgmL-1. Regarding the SAB concentrations, 5 μgmL^−1^ and 50 μgmL^−1^ were diluted in PBS (1:1000 and 1:100 ratios) and used to identify the concentration at which the surface was coated with IgG without compromising the interaction with the antigen.

For the experiments, we employ a Surface Plasmon Resonance (SPR) instrument that operates in angular interrogation mode (AIM). The instrument comprises a red laser light source, lenses for optical beam coupling, an image detector, and electronic components [18]. Figure 1 depicts the schematic diagram of the SPR sensor. The SPR phenomenon refers to the coupling between electromagnetic and surface plasmon waves, on the interface of two different media with different dielectric constants (normally, a metal film and a dielectric). When the light excitation condition is fixed, the SPR technique allows precise measurement of changes in the: 1) refractive index; 2) thickness of the medium adjacent to the metal film; and 3) adsorption layer on the metal surface [19, 20]. In AIM, a significant loss in reflected light at a specific angle for a fixed light source wavelength is observed. As the venom material diffuses out of the serum solution, the SPR instrument indicates the shift in the angular position of the intensity minimum. Here, it is represented as a variation of the effective refractive index (RIU), displayed as a so-called SPR-sensorgram.

**Figure 1.**
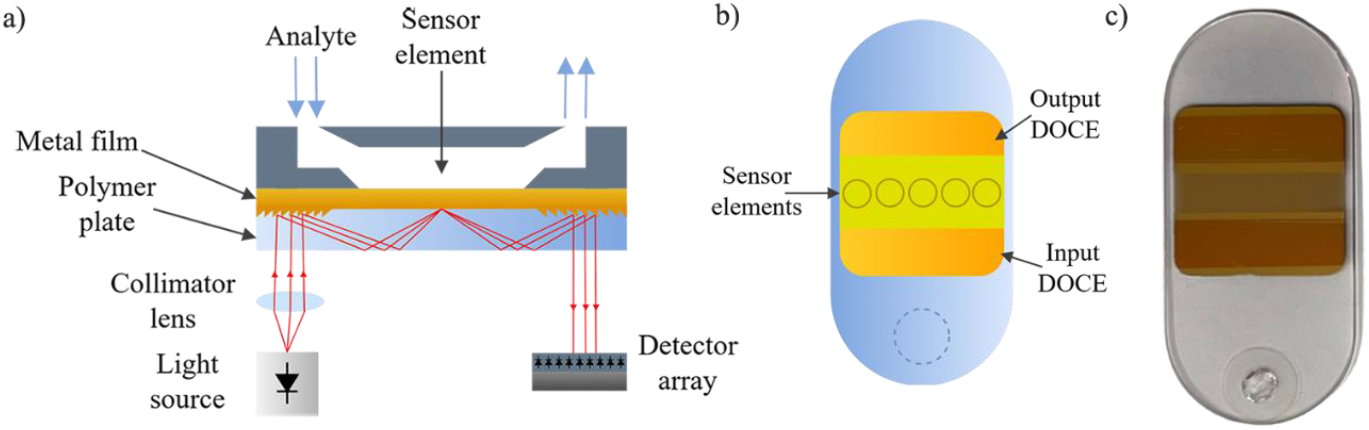
Schematic (not in scale) of SPR instrument set-up (a) with an exchangeable polymer chip with optical grating couplers, input DOCE and output DOCE (b), and the photograph of used chip (c).

Figure 2 shows a representation of the arrangement of the layers formed, which follows the following order of addition: first the antivenom serum (containing antibodies), followed by the *Crotalus* snake venom and, lastly, the Bothrops snake venom (to assess the presence or absence of cross-reaction between the antibothrops serum and the Crotalus venom simultaneously with the test to assess the specificity between the antibothrops serum and the Bothrops venom).

**Figure 2.**
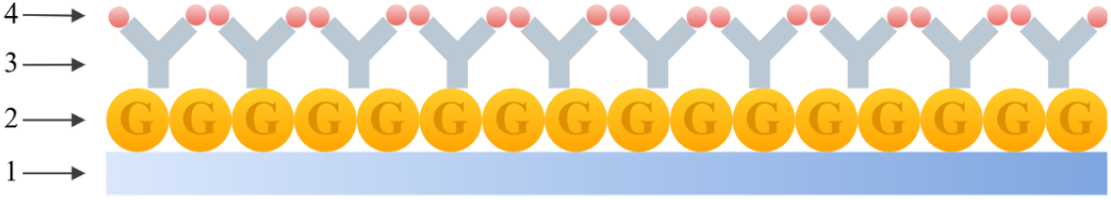
Representation of the arrangement of the layers: 1. polymer substrate with a 2. thin 50 nm gold layer (G, in yellow); 3. antibodies (Y, in gray); 4. Snake venom (spheres in red). The solutions were transported to the biochip by a peristaltic pump. Flow is essential due to the need to measure multiple analytes in a single assay. The flow rate varies between 5 and 10μL, values directly related to the analyte, to optimize the adsorption and desorption process of the analyte on the gold film.

Initially, the gold surface of the biochip was washed with deionized water while the biochip was outside the sensor, followed by a second wash with deionized water after coupling the biochip to the sensor to avoid any possible air contamination, as well as to have a reading that started at zero, since deionized water was used as a reference. From the moment the water flow entered, the signal began to be measured and recorded. The flow was paused for a few seconds so that the input channel could be manually transferred to the next solution. The graphs were generated when the interactions on the sensor surface occurred.

Figure 3 illustrates how the results are presented, following the steps described below.

**Figure 3.**
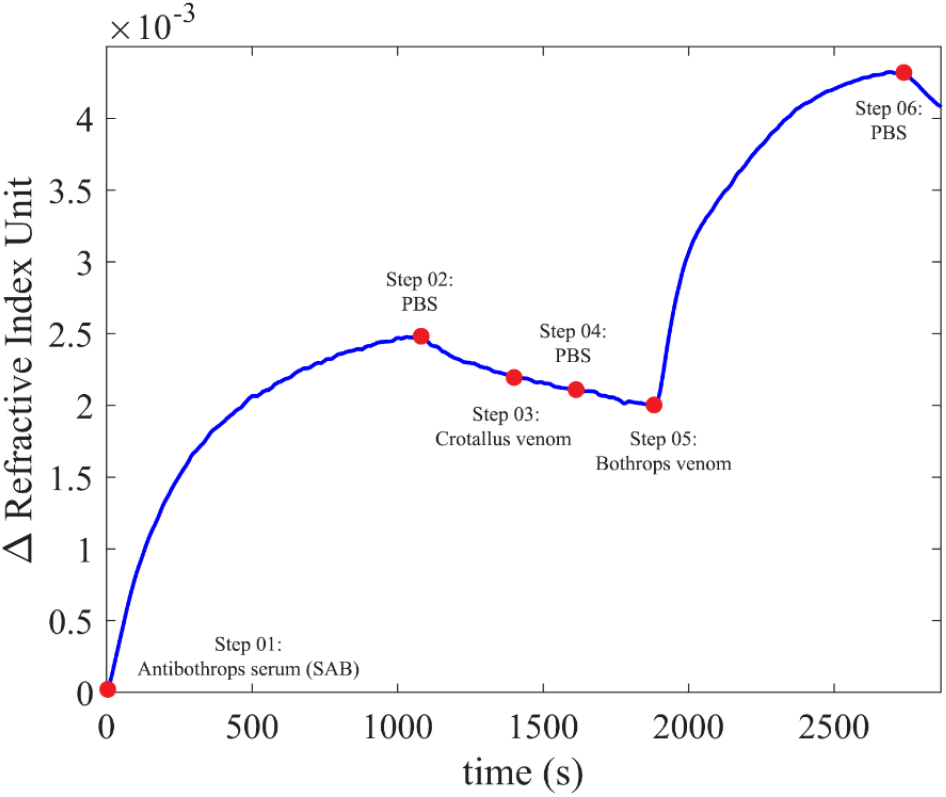
Illustrative graph demonstrating the test stages that make up the results. The blue markers on the graph indicate the time point at which the type of fluid in the pump reservoir was changed.

Step 1: the anti-bothrops serum (SAB) diluted in PBS was admitted; the physical adsorption of the antibodies on the gold was evidenced by the increase in the line on the graph, and when the line became constant (indicating total coverage of the film) the flow was paused.

Step 2: the inlet channel was transferred to the PBS solution, so that the weakly bound molecules were removed (desorption process), an event indicated by the decay of the line on the graph; the flow was paused.

Step 3: the inlet channel was transferred to the Crotalus venom solution diluted in PBS; after the line on the graph became constant, the flow was paused.

Step 4: PBS was admitted again, and, after the line on the graph remained constant, the flow was paused.

Step 5: the inlet channel was transferred to the Bothrops venom solution diluted in PBS; when the line on the graph became constant, the flow was stopped.

Step 6: the inlet channel was transferred to the PBS solution so that the molecules (present in the venom) weakly bound to the previous layer were removed.

At the end of each experiment, a cleaning cycle was performed to remove the substances adsorbed on the sensor surface. This consisted of flowing deionized water over the surface, followed by enzymatic detergent (Extran) diluted in deionized water, and then deionized water again. This washing procedure did not cause any damage to the gold film. After the surface cleaning step, a new experiment could be performed using the same chip. All tests were performed in triplicate.

The experiments were performed at the Biosensors Laboratory of the Department of Electrical Engineering (DEE) - Center for Electrical and Computer Engineering (CEEI), at the Federal University of Campina Grande (UFCG), located in the state of Paraíba, Brazil.

To analyze kinetic interactions, we apply an approach based on the one-to-one Langmuir binding model, which assumes homogeneous binding sites and no mass transport limitations. Considering the interaction between the sensor chip surface (G) and the injected analyte (A) following the one-to-one Langmuir model for complex formation (AG) in a flow-cell-based SPR system, continuous flow maintains A constant. Therefore, during analyte injection phase, A is equal to C_A_, and during the buffer (PBS) injection phase, A is equal to zero. The sensor response varies with time according to the ordinary differential equation [21, 22]:

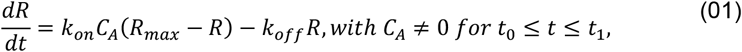

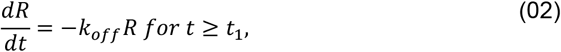

where, *R* is the observed response and *R*_*max*_ corresponds to the maximal response that would be observed if an infinite concentration of A was injected. *k*_*on*_ represents the association constant and *k*_*off*_ the dissociation constant.

By analytically solving the ordinary equation under the assumption of homogenous binding sites. If no mobile analyte has been initially bound to the gold surface, the time course of association is described by an exponential equation [23]:

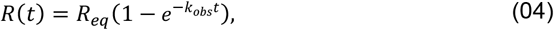

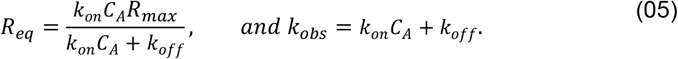

If the free analyte is removed from the buffer (*t ≥ t*_*1*_), the complex AG dissociates exponentially with time as expressed in the equation [23]:

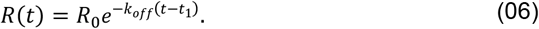

in which, *R*_*0*_ represents the sensor response at the start of dissociation. In this model, the observed binding rate constant *k*_*obs*_ in the association phase is always higher than the dissociation rate constant *k*_*off*_.

In our analysis, the model parameters θ = [*R*_*eq*,_ *k*_*obs*,_] for association phase and θ = [*R*_*0*,_ *k*_*off*,_] for dissociation phase and were optimized by minimizing the sum of squared residuals (SSR) between the experimental data *R(t)* and the model prediction *R*^*\**^*(t; θ)* as follows:

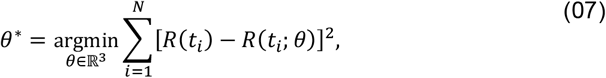

where, *N* is the number of datapoints. The constrained nonlinear least squares problem was solved using the trust-region reflective least squares algorithm [24,25]. The algorithm adopted a termination tolerance on the function value of *10*^*-6*^ with a termination tolerance on θ of *10*^*-6*^. We consider as constraints the lower and upper bounds given by 0 < *R*_*eq*_ < ∞ and 0 < *k*_*obs*_ < ∞. As initial guesses, we considered θ = [*max(R(t))*, *0*.*1*] with *t*_*0*_ *≤ t ≤ t*_*1*_ for association phase and θ = [*R(t=1)*, *10*^*-3*^] with *t ≥ t*_*1*_ for dissociation phase.

## 5 RESULTS

The interaction between antibodies and antigens was evaluated at different concentrations of venom and antivenom, by the change in the refractive index for each substance (Figures 4 and 5). In the experiments, the binding of SAB to the sensor surface and the binding capacity of venom to the IgG layer were investigated at two different concentrations of SAB. The results of the interaction between antibodies and the corresponding (Bothropic) and non-corresponding (Crotalus) venom are shown in Figures 4 and 5.

**Figure 4.**
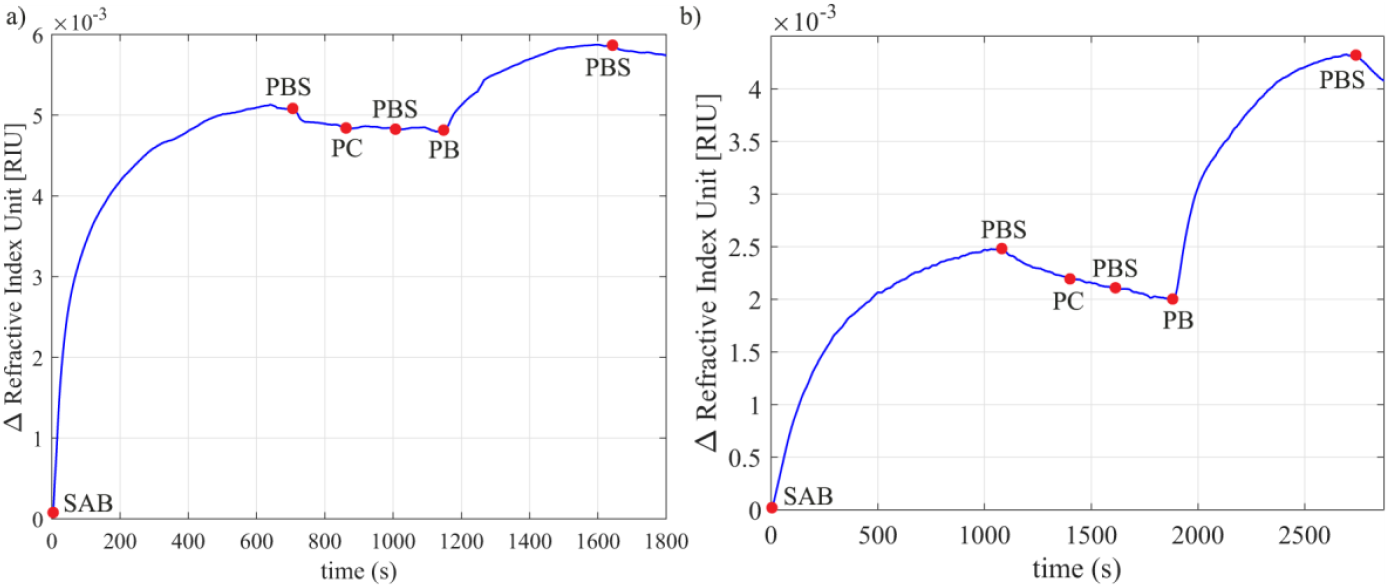
Graphs of the moving average of the refractive indices of PBS, Bothrops Antivenom (SAB), Crotalus Venom (PC) and Bothrops Venom (PB) as a function of time. a) SAB 1:100, PC 1:7,500, PB 1:7,500; b) SAB 1:1,000, PC 1:7,500, PB 1:7,500. The blue markers on the graph indicate the time point at which the type of fluid in the pump reservoir was changed.

**Figure 5.**
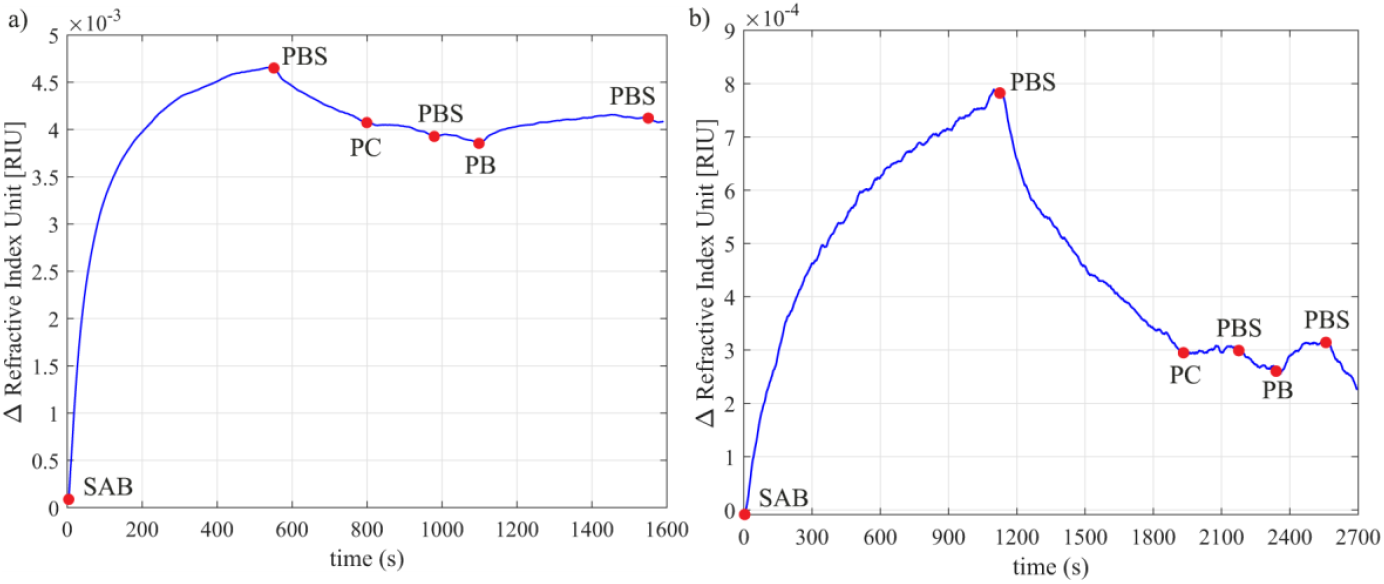
Graphs of the moving average of the refractive indices of PBS, Bothrops Antivenom (SAB), Crotalus Venom (PC) and Bothrops Venom (PB) as a function of time. a) SAB 1:100, PC 1:60,000, PB 1:60,000; b) SAB 1:1,000, PC 1:60,000, PB 1:60,000. The blue markers on the graph indicate the time point at which the type of fluid in the pump reservoir was changed.

Graphs of the moving average of the refractive indices of PBS, Bothrops Antivenom (SAB), Crotalus Venom (PC) and Bothrops Venom (PB) as a function of time. A: SAB 1:100, PC 1:60,000, PB 1:60,000; B: SAB 1:1,000, PC 1:60,000, PB 1:60,000. The blue markers on the graph indicate the time point at which the type of fluid in the pump reservoir was changed.

It is possible to observe the change in the sensor response as the antibodies and antigens interact, as shown in the curves shown in Figures 4 and Each curve corresponds to the adsorption/desorption of a specific substance, showing the deposition of layers on the sensor surface, confirming the conjugation between bioreceptor and analyte. The sensor signal tends to return to the baseline when the flow containing venom is interrupted and exchanged for PBS, due to the dissociation process. Although at a concentration of 1:60.000, Bothrops venom dissociates more easily from antibodies due to its low concentration, the sensor detected the adsorption of this venom.

The intensity of the sensor response to venom was proportional to the concentrations of venom and IgG (SAB) used. The highest venom concentration (6.784 μgmL^−1^) generated a higher response when using the lowest IgG concentration (5 μgmL^−1^), on the other hand, when using the highest IgG concentration (50 μgmL^−1^), the refractive index of the venom at high concentration was lower. When we tested the lowest venom concentration (0.848 μgmL^−1^) together with the lowest IgG concentration, there was a higher refractive index of the venom; at the high IgG concentration the refractive index of the venom was lower. From Figures 3A and 4A, it is possible to observe that as the venom protein concentration increased from 0.848 μgmL^−1^ (venoms in a ratio of 1:60.000) to 6.784 μgmL^−1^ (venoms in a ratio of 1:7.500), the refractive index increased by 3.91, compared to immunoglobulins at a concentration of 5 μgmL^−1^. Similarly, Figures 4B and 5B show that as the venom protein concentration increased from 0.848 μgmL^−1^ to 6.784 μgmL^−1^, the refractive index increased by 0.75, compared to immunoglobulins at a concentration of 50 μgmL^−1^. Regarding the biosensor response time, the final time of the tests varied between 25 and 40 minutes.

Figures 4 and 5 also reveal the absence of a variation in the refractive index of Crotalus venom on Bothrops antibodies, thus confirming the absence of cross-interaction. We analyzed the adsorption of IgG (SAB) by implementing curve fitting to estimate the association and dissociation constants. The experimental data (represented by the blue line) is plotted alongside the estimated data (shown as a dashed green line) for the experiments in Figure 6.

**Figure 6.**
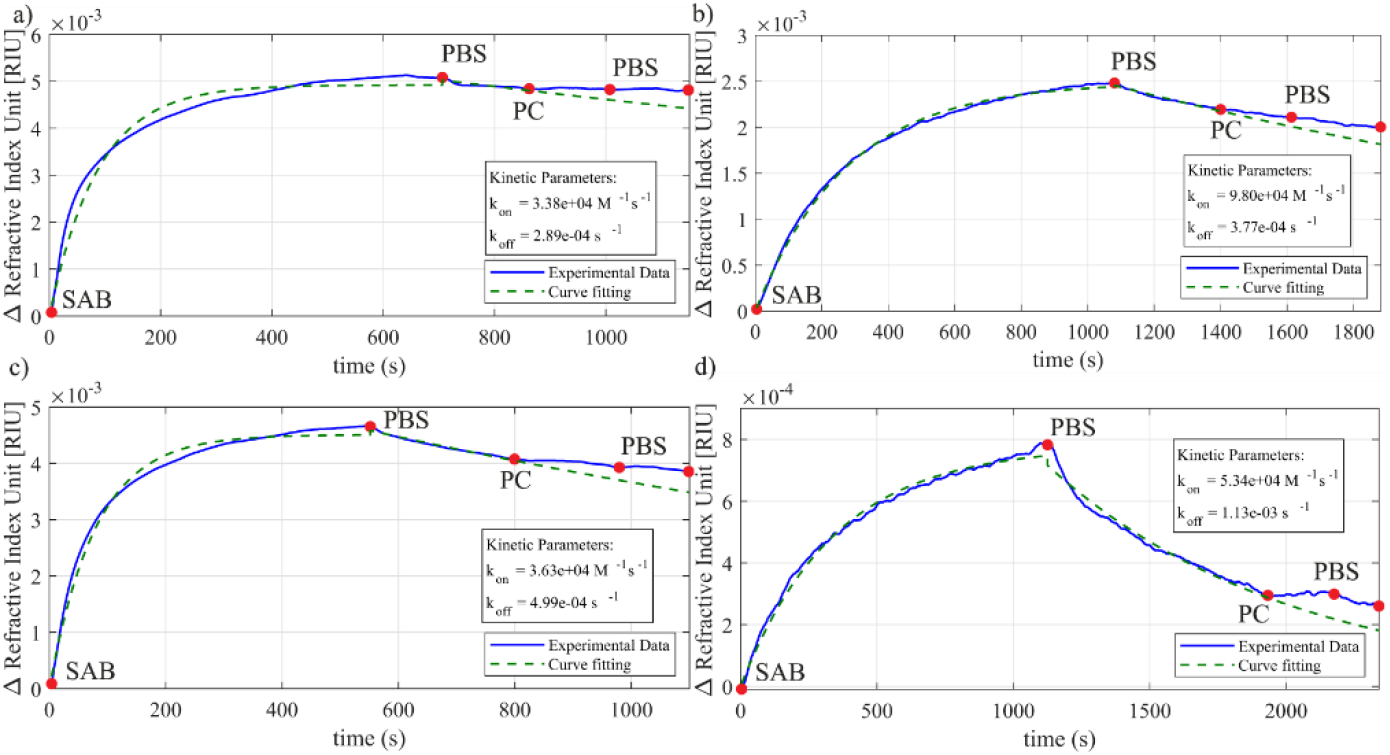
Kinetics constant estimation for different concentration of Bothrops Antivenom (SAB): a) SAB 1:100, PC 1:7,500, PB 1:7,500; b) SAB 1:1.000, PC 1:7,500, PB 1:7,500; c) SAB 1:100, PC 1:60,000, PB 1:60,000; d) SAB 1:1.000, PC 1:60,000, PB 1:60,000.

To estimate the kinetics parameters, we considered the association time as the time comprised in the interval between the red dot SAB and the first red dot PBS. The dissociation time corresponds to the interval between the first red dot PBS and PC injection. After estimating the parameters, we apply the values in equations 04 and 06, covering the dissociation interval from the first PBS injection to the end of the second PBS injection cycle. According to the estimated curve (dashed green line), we would have a higher quantity of SAB removed from the sensor surface by injecting only PBS instead of PC. However, the second PBS cycle removed all possible molecules in the PC solution, approximating the sensor response to the estimated dissociation curve and showing the specificity of SAB.

To confirm the selectivity between Bothrops antibodies (SAB) and Bothrops venom, Bothrops venom was promptly added after the passage of the Crotalus venom (both venoms at the same concentration). With the presence of Bothrops venom, the occurrence of interaction between the SAB and Bothrops venom was observed, thus, the previous passage of the fluid containing the Crotalus venom did not affect the interaction between the corresponding antibodies and venom. Which means that, even when submitted first to the system, the Crotalus venom is not capable to interact or modify the link between Bothrops venom and its specific antivenom. Which could happen as venoms share similar epitopes [26] and can elicit cross reactions [27]. This data shows that the SPR sensor allied to the simple methodology developed, is capable to go forward and be used to discriminate between these two venoms at point of care institutions.

## 6 DISCUSSION

In this article, we report a simple methodology for discriminate between *Bothrops* and *Crotalus* venoms using a surface plasmon resonance biosensor, based on the evaluation of antigen-antibody interaction. To more accurately reproduce the real situation, we used antivenom as a bioreceptor, since it is the most common and accessible form of obtaining antibodies in health centers. We also used crude snake venom, considering that this is the form in which venom can be found in body fluids available for evaluation, such as blood and urine.

Two groups of tests (Figures 4 and 5) were performed; in each group a specific concentration of antibodies was used and the venom concentration varied. It was observed that the refractive index of the antibody layer was different for each test in which the same concentration of SAB was used. This difference in response, even using the same concentration of SAB, can be attributed to the packaging of antibodies [28], which, due to immobilization by physical adsorption, can occur in different ways, since in this type of adsorption the antibodies spontaneously bind to the surface of the sensor, causing a disordered orientation [29, 30, 31].

Figures 4 and 5 reveal that the presence of high venom concentrations results in a greater shift in the resonance angle compared to tests in which low venom concentrations were applied (which resulted in a smaller shift in the resonance angle). Thus, the existence of a direct relationship between the concentration of the substance and the intensity of the response is demonstrated [32, 33].

Two different concentrations of IgG were used to evaluate the intensity of venom binding; thus, a relationship was observed between the concentration of antibodies and the amplitude of the venom response. When the antibody solution was less concentrated, the venom at a high concentration elicited a more intense response. In contrast, in the experiments in which the antibody solution was more concentrated, the venom at a high concentration resulted in a less pronounced response. High surface coating densities have been shown to reduce antibody binding capacity due to spatial constraints, as antibodies obstruct each other’s access to binding sites [34]. Thus, at a lower concentration, the binding capacity between the IgG domain and the antigen is increased.

An important attribute observed was the reusability of the chip used: after each test was completed, the surface cleaning procedure to remove the adsorbed substances proved to be efficient, since the signal returned completely to its baseline, established by passing a flow of deionized water. Thus, subsequent measurements were not affected by the later measurements. This data is significant because underlines the importance of reusability of this type of system on a point of care institution, even more if it is supported by public financial incentive.

Regarding the composition of snake venoms, it is important to highlight that the diversity of their composition is present at all hierarchical levels of snake taxonomy; the complex variability is the result of genetic mutations, genetic drift and natural selection, thus, the venom components have adjusted to the specific needs of each group of snakes [35, 36]. However, despite the evolutionary changes in the genomic and proteomic composition of venoms, some families of toxins are present in the three families of snakes that have frontal fangs, since the venom glands of snakes are homologous [26]. The omnipresence of certain toxins in different snake families leads to the possibility of cross-reaction between the antivenom and the non-corresponding venom at the time of detection.

However, it is possible to observe in the graphs in Figures 4 and 5 that there was no shift in the curve indicative of interaction between the Bothrops antibodies (SAB) and the Crotalus venom. Thus, our tests confirmed the selectivity between the bothropic antibodies and the Bothrops venom. Similarly, a previous study developing an impedimetric immunosensor for the diagnosis of snake envenomation did not report significant variations indicating the formation of the immunocomplex between the Bothrops antibody and Crotalus venom, even at high concentrations [33].

The coexistence of toxins in different species has enabled the production of antivenom with the potential to neutralize venoms from different snake species, promoting the production of broad-spectrum antivenoms using a mixture of crude venoms and toxin fractions [37], as is the case of the Brazilian Bothrops antivenom, generated through the venom of five species (B. jararaca, B. alternatus, B. jararacussu, B. moojeni and B. neuwiedi) [16]. It is possible to observe that the venom of the species B. erythromelas is not included in the pool used in the production of the antivenom. Even so, the biosensor used in this research detected a clear interaction between the venom and the antivenom, even at low concentrations. This occurs due to the sharing of certain toxins among species of the same genus [26]. However, the addition of B. erythromelas venom in the production of brazilian bothrops antivenom could amplify detection by the biosensor, considering that there would be a higher response due to the interaction of B. erythromelas venom with antibodies specific to toxins of this species. This observation is corroborated by studies that identified components of B. erythromelas not neutralized by Brazilian Bothrops antivenom [37, 38, 39]. It is important to emphasize the need to expand the diversity of Bothrops snake venoms intended to produce Bothrops antivenom, considering that snakebites in Brazil are predominantly caused by snakes of this genus [4].

A surface plasmon resonance (SPR) sensor was developed for the detection of crude venom from the Indian cobra (*Naja naja*) and a polyvalent antivenom [40]. As in this work, all measurements were performed in phosphate-buffered saline solution and the implication of other components existing in the blood was not evaluated. After testing different antibody concentrations and different incubation times, all sensors were constructed using a concentration of 2mg/ml of antibody incubated for 8 hours, and the venom diluted in PBS was exposed to the sensor for 12 minutes. The sensor detected the venom at concentrations between 0.1 mgmL^−1^ and 1.0 mgmL^−1^. Although the authors used the same class of biosensors (SPR) and a methodology similar, the biosensor used in our research showed greater sensitivity to the corresponding antigen-antibody interaction, given that it detected the venom at a significantly lower concentration (0.848 μgmL^−1^), thereby reinforcing the relevance of the proposed technology and methodology for advancing the implementation of a diagnostic tool for snake envenoming.

## Data Availability

UPON REQUEST

## ACKNOWLEDGMENTS

This work was partially funded by the project “SmartSens Base Systems,” supported by the EMBRAPII VIRTUS Competence Center in Intelligent Hardware for Industry – VIRTUS-CC (MCTI grant number 055/2023), the Paraíba State Research Foundation (FAPESQ, grant number 09/2023), and in part by the Coordenação de Aperfeiçoamento de Pessoal de Nível Superior – Brasil (CAPES), Finance Code 001. The authors would also like to thank the Department of Electrical Engineering (DEE/UFCG) and the Center for Electrical Engineering and Informatics (CEEI) at the Federal University of Campina Grande (UFCG) for providing the research infrastructure used during this investigation.

